# COVID-19 ASSOCIATED MUCORMYCOSIS: A CASE-CONTROL STUDY

**DOI:** 10.1101/2021.08.16.21262109

**Authors:** Dulari Gupta, Rahul Kulkarni, Shripad Pujari, Atul Mulay

**Affiliations:** Deenanath Mangeshkar Hospital and Research Center, Pune, Maharashtra, India 411004

**Author notes:** Corresponding Author: Dr. Atul V. Mulay, Address: Deenanath Mangeshkar Hospital and Research Center, Erandawane, Pune, Maharashtra, India. 411004, Email address.

**Keywords:** Covid19, Mucormycosis, Case-control study, Steroids, Diabetes

## Abstract

**Background:** India has seen a surge in COVID-19 associated mucormycosis (CAM) cases during the second wave of the pandemic. We conducted a study to determine independent risk factors for CAM.

**Methods:** We performed a retrospective case control study in a tertiary care private hospital in Pune, India. Fifty-two cases of CAM were compared with 166 concurrent controls randomly selected from the COVID-19 admissions during the same time period. Association of demographic factors, comorbidities, cumulative steroid dose used (calculated as dexamethasone equivalent), maximum respiratory support required, use of injectable/oral anticoagulation, and use of aspirin with CAM was assessed by univariate and multivariate logistic regression.

**Results:** A total of 218 subjects (52 cases; 166 controls) were studied. Any diabetes (pre-existing diabetes and new onset diabetes during COVID-19) was noted in a significantly higher proportion of cases (73·1%, 45·8% P<0.001) and cumulative dexamethasone dose used in cases was significantly greater (97·72 mg vs 60 mg; P=0·016). In a multivariate regression analysis cumulate dexamethasone dose >120 mg (OR 9·03, confidence interval 1·75-46·59, P=0·009) and any diabetes (OR 4·78, confidence interval 1·46-15·65, P=0·01) were found to be risk factors for CAM. While use of anticoagulation (OR 0·01, confidence interval 0·00-0·09, P<0·001) and use of aspirin (OR 0·02, confidence interval 0·01-0·07, P<0·001) were found to be protective against CAM.

**Conclusion:** Diabetes mellitus and cumulative dose of dexamethasone greater than 120 mg (or equivalent dose of other corticosteroid) were associated with an increased risk of CAM while use of aspirin and anticoagulation were associated with a lower risk.

## Introduction

Coronavirus disease (COVID-19) is a disease caused by severe acute respiratory syndrome coronavirus 2 (SARS-CoV-2), which was declared as a pandemic by World Health Organization (WHO) in March 2020 ^1^. India has the second highest burden of COVID-19 cases, next only to United States ^2^. First wave of COVID-19 in India peaked in the middle of September 2020 and cases declined by end of December 2020. Second wave of COVID-19 started in India in February 2021, peaked by May 2021 and is now declining ^3^. India recorded around 10 million cases in the first wave and an additional 20 million cases in the second wave until second half of June 2021.

Bacterial and fungal co-infections are seen in up to 8% patients with COVID-19 ^4^. An unusual global observation during the course of the second wave was a large number of mucormycosis cases associated with COVID-19 out of which more than 80% were reported from India ^5–8^. According to various reports, till May 28 2021 14872 cases of CAM ^9^ and until 3^rd^ June 2021, more than 31000 cases of COVID-19 associated mucormycosis (CAM) were reported from the country ^10^.

Mucormycosis is an invasive fungal infection caused by the order *Mucorales*. Mucormycosis can spread to humans through three routes: inhalation of spores, ingestion of spores in food and traumatic inoculation. These fungi are ubiquitous in the environment and are seen microscopically as broad septate or aseptate right angled branching fungi ^11^. According to Patel et al diabetes, diabetic ketoacidosis, organ transplant, malignancy, corticosteroids, immunosuppression, trauma, and burns are known risk factors for mucormycosis ^12^. India has the second largest population of diabetics in the world ^13^. Corticosteroids in general and dexamethasone in particular have been recommended for the treatment for severe COVID-19^14^. COVID-19 by itself has also been shown to be associated with new onset diabetes mellitus^15^. India has a high background incidence of non-COVID-19 mucormycosis - 140 cases per million population which is 70 times higher than global incidence ^16,17^. These factors may partly account for increased reporting of CAM in India.

Among the Indian states, Maharashtra is the worst affected by COVID 19 (17). It is also the state with the second highest number of cases of CAM ^18^. Till 14^th^ July Maharashtra recorded 9268 cases of CAM (incidence less than 0.3%) and 1112 deaths due to CAM ^19^. Incidence of CAM was estimated to be 0·27% (0·05% to 0·57% in individual centers) in a multicenter study from seven centers ^12^. CAM therefore appears to be a rare complication during the course of COVID-19. Earlier studies have implicated oxygen use (57%); diabetes mellitus (78%) and corticosteroid use (87%) in the etiology of COVID associated mucormycosis ^18^. Earlier studies were mostly case series without a control group. We undertook a case-control study to determine the risk factors of CAM, in particular to further elucidate role of corticosteroid dose that may increase risk of CAM.

## Methods

We conducted a case control study at an 800 bedded tertiary level referral hospital in Pune, Maharashtra, India. The study was approved by the Institutional Ethics Committee.

### Selection of cases

We defined cases as patients admitted in our hospital for treatment of COVID-19 associated rhino-orbito-cerebral mucormycosis (ROCM). To be included in this study as cases, the following criteria had to be fulfilled: 1. Age >18 years, of either sex; 2. Diagnosis of mucormycosis had to be confirmed by laboratory demonstration of mucor in specimens by staining and/or culture and/or histopathology; 3. Mucormycosis was diagnosed within 60 days of prior diagnosis of COVID-19 ^20,21^; 4. Patients were admitted in our hospital for treatment of mucormycosis from 1^st^ February 2021 to 31^st^ May 2021.

### Selection of controls

A total of 3101 patients with COVID-19 were admitted in our hospital during the study period. Controls were patients with COVID-19 disease, confirmed by Rt-PCR or rapid antigen test, age > 18 years of either sex, and who did not develop invasive fungal disease within 60 days after diagnosis of COVID-19 (all included controls were telephonically contacted to ascertain that they did not develop mucormycosis or any other serious illness requiring hospital admission within 60 days after discharge). For each case included in the study, we prepared a list of COVID-19 patients admitted to the hospital within seven days of onset of COVID-19 of the index case. From this list, three random controls were selected for each patient with mucormycosis using randomly generated numbers. Patients were excluded from the control group if 1) they had received treatment for COVID-19 at other hospitals prior to admission or 2) they died with COVID-19 within 7 days of hospitalization or 3) they were admitted for some other primary disease, and later incidentally found to be RT PCR positive for COVID-19. Though it would have been ideal to select controls from the hospitals which referred the patients with mucormycosis, it was not possible since accurate records from the referring hospitals were not available.

We prepared a standardized form (appendix 1) to extract data from in-patient case records of cases and controls. The extracted data included age, sex, duration of hospitalization for COIVD-19, comorbidities (diabetes mellitus, systemic hypertension, chronic kidney disease, new onset diabetes after COVID-19, underlying malignancy, chronic immune-suppression, and any other relevant medical condition), use of corticosteroids, remdesivir, tocilizumab, low molecular weight heparin (LMWH), aspirin, newer oral anticoagulants (NOAC), cumulative corticosteroid dose used and maximum respiratory support required (no oxygen, oxygen via nasal canula or face mask, non-invasive ventilation or invasive mechanical ventilation) and laboratory parameters.

### Statistical Analysis

We describe baseline characteristics as mean (±SD) or median (inter quartile range) as appropriate for continuous variables and proportions for categorical variables. We compared continuous variables between cases and controls using Students’ t-test or Mann-Whittney U test as appropriate. We compared categorical variables using chi square test or Fisher’s exact test as appropriate.

We determined independent risk factors associated with mucormycosis by multivariate logistic regression. Main risk factors of interest were cumulative corticosteroid dose and use of tocilizumab for COVID-19. Where corticosteroids other than dexamethasone (methylprednisolone, prednisolone, hydrocortisone) were used, we converted the corticosteroid dose into dexamethasone equivalents. The cumulative corticosteroid dose was categorized as less than 60 mg (dexamethasone equivalent), 60-120 mg and more than 120 mg according to the recommended dexamethasone dose of 6 mg per day for a maximum of 10 days by the RECOVERY trial ^14^. We considered severity of COVID-19, age, sex, diabetes mellitus, hypertension, chronic kidney disease, immune-compromised status, and use of aspirin, low molecular weight heparin, NOAC as potential confounders for inclusion in multivariate model. COVID-19 severity was classified according to WHO criteria^22^: mild (not requiring oxygen or admission to hospital), moderate (requiring hospitalization but no supplemental oxygen), severe (requiring supplemental oxygen), and critical (requiring non-invasive ventilation or mechanical ventilation). Variables with a p value <0.25 in univariate analysis, were included in the multivariate logistic regression analysis and final model was selected by backward elimination. In addition, in a sensitivity analysis, we tested association of cumulative corticosteroid dose of greater than 120 mg dexamethasone equivalent with the risk of mucormycosis by propensity score covariate adjustment. We used age, sex, COVID-19 severity, C-reactive-protein (CRP) level and neutrophil to lymphocyte ratio in a logistic regression model to calculate probability of receiving cumulative steroid dose >120 mg dexamethasone equivalent. For all analyses, we considered two-sided p value <0·05 as statistically significant. All statistical analyses were performed using SPSS version 26 (SPSS for Windows, Chicago, SPSS Inc).

## Results

During the study period, 101 patients suspected to have COVID-19 associated ROCM were admitted to our hospital. Of these, 5 did not have demonstrable mucor in clinical specimen and in 44 patients details of previous COVID-19 treatment were not available. Thus, we included 52 patients of mucormycosis for the final analysis. A list of 197 controls was randomly generated as described earlier. Of these, 29 died within seven days of hospitalization and 2 developed mucormycosis within 60 days of onset of COVID-19 and were excluded. We, therefore had 166 controls for the 52 patients with mucormycosis.

Baseline characteristics of the cases and controls are shown in Table 1. The majority of patients were males in the 5^th^ decade of life. It is noteworthy that very few patients had chronic kidney disease or immunocompromised state at baseline. The severity of COVID-19 was moderate in 12/52 (23·1%) patients, severe in 32/52 (62·7%) patients and critical in 7/52 (13·7%) patients. Median duration of hospitalization for COVID-19 was 9 (IQR 6-12·25) days for the cases and 6 (IQR 4-8) days for the controls (p=0·0001). Median time from onset of COVID-19 to diagnosis of mucormycosis was 14 days (IQR 10-20 days). Select laboratory parameters and treatment details of cases and controls are given in Table 2.

**Table 1:** Demographic characteristics of cases and controls.

|  | Cases (N=52) | Controls (N=166) | P |
| --- | --- | --- | --- |
| Age | Median 53.38 (42.25-62.75) | Median 55.50 (44.75-68.25) | 0.236 |
| Gender (male) | 41 (78.8%) | 110 (66.3%) | 0.086 |
| Pre-existing DM | 27 (51.9%) | 59 (35.5%) | 0.035 |
| HT | 22 (42.3%) | 66 (39.8%) | 0.74 |
| CKD | 6 (11.5%) | 8 (4.8%) | 0.11 |
| Chronic Immunosuppression | 0 | 3 (1.8%) | 1 |
| Newly diagnosed DM | 11 (21.2%) | 17 (10.2%) | 0.04 |
| Any DM | 38 (73.1%) | 76 (45.8%) | 0.001 |

(DM- diabetes mellitus, HT-hypertension, CKD- chronic kidney disease)

**Table 2:**
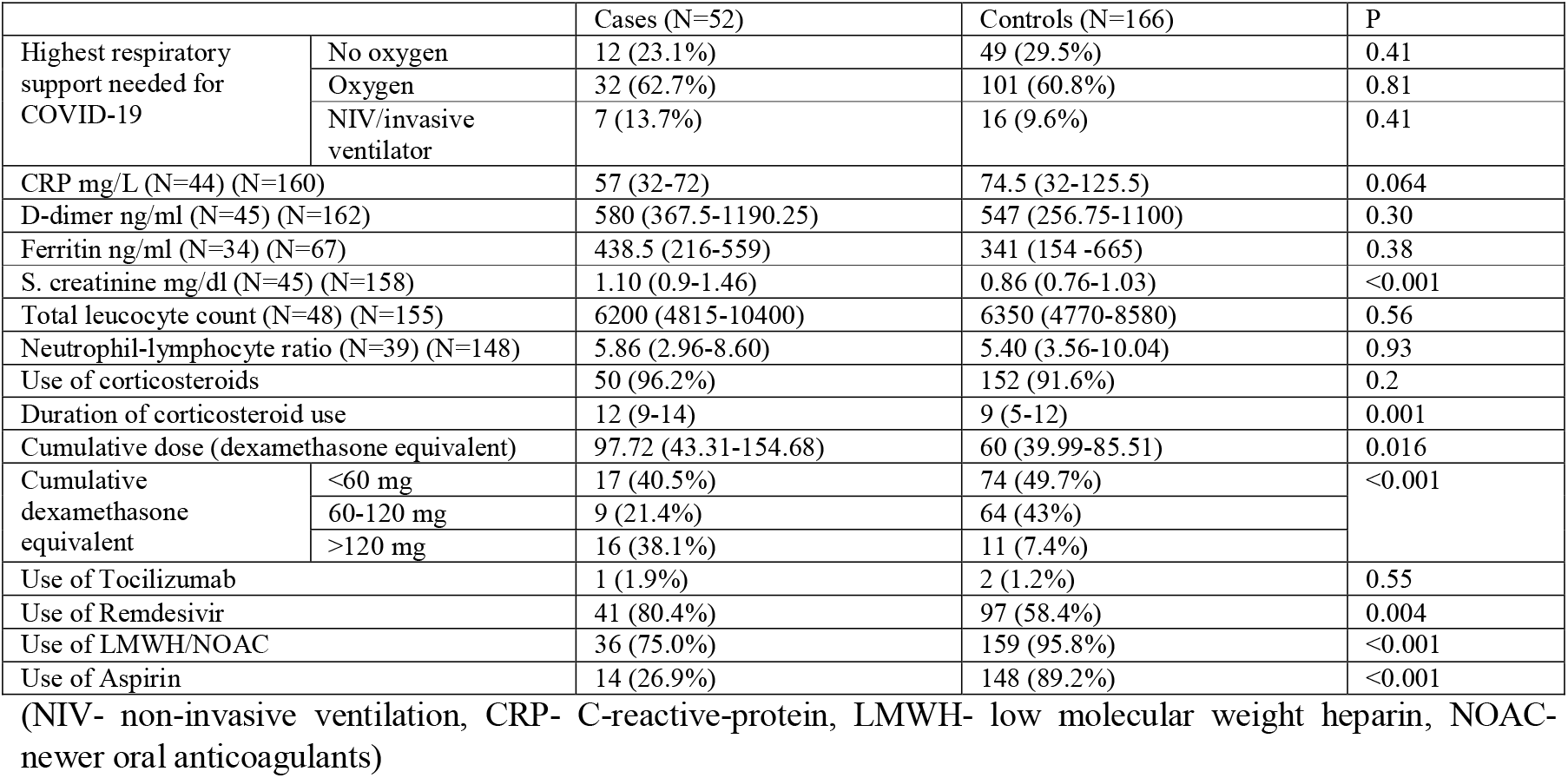
Characteristics of COVID-19 and treatment of COVID-19.

Mucormycosis was diagnosed on the basis of stained smears (Potassium hydroxide and Calcofluor) in 45/52 (86·8%); histopathology in 42/52 (80·7%) and positive fungal cultures in 25/52 (48%) cases. Eleven cases (21·1%) had dual infection with both mucormycosis and aspergillosis. The presentation of fungal infection was headache 48/52 (92·3%), facial pain 42/52 (80·8%), ophthalmoplegia 14/52 (26·9%), facial numbness 13/52 (25%), loss of vision 12/52 (23·1%), focal neurological deficits 4/52 (7·7%) and loosening of teeth 2/52 (3·8%).

In the univariate logistic regression, we found that presence of diabetes and high corticosteroid dose were significantly associated with an increased risk of mucormycosis whereas aspirin and anticoagulation were associated with a significantly decreased risk. In a multivariate logistic regression model, cumulative steroid dose greater than 120 mg dexamethasone equivalent (OR 9.03, 95% confidence interval 1.75-46.59, P=0.009) and presence of diabetes (OR 4.78, 95% confidence interval 1.46 – 15.65, P=0.001) were associated with a significant independent increased risk of mucormycosis while aspirin (OR 0.02, 95% confidence interval 0.01-0.07, P<0.001) and anticoagulation (OR 0.01, 95% confidence interval 0.00-0.09, P<0.001) were associated with a significant decreased risk. (Table 3)

**Table 3:** Independent determinants of COVID-19 associated mucormycosis (Multivariate Logistic Regression)

|  | Odds ratio | 95% confidence Interval | P |
| --- | --- | --- | --- |
| Cumulative dexamethasone dose <60 mg | Ref |  |  |
| Cumulative dexamethasone dose 60-120 mg | 1.01 | 0.29-3.57 | 0.986 |
| Cumulative dexamethasone dose >120 mg | 9.03 | 1.75-46.59 | 0.009 |
| Any Diabetes mellitus | 4.78 | 1.46-15.65 | 0.010 |
| Use of Aspirin | 0.02 | 0.01-0.07 | <0.001 |
| Use of Anticoagulation | 0.01 | 0.00-0.09 | <0.001 |
| No supplemental oxygen | Ref |  |  |
| Supplemental oxygen | 3.75 | 0.86-6.29 | 0.078 |
| Mechanical respiratory support (non-invasive or invasive ventilation) | 2.14 | 0.26-17.80 | 0.480 |

In a sensitivity analysis, a propensity score covariate adjusted model did not change association of high corticosteroid dose with mucormycosis. Inclusion of remdesivir in the multivariate model also did not change these findings. A similar regression analysis without exclusion of those controls who developed mucormycosis later, also did not change the results of the regression model.

## Discussion

In this case control study of COVID-19 associated mucormycosis (CAM), we demonstrate that cumulative corticosteroid dose more than 120 mg dexamethasone equivalent and presence of diabetes mellitus (either known diabetes or diabetes that was detected for the first-time during admission for COVID-19) are factors associated with an independent increased risk of mucormycosis, whereas use of aspirin and therapeutic anticoagulation were negatively associated with risk of mucormycosis.

Diabetes mellitus, particularly with ketoacidosis is the commonest reported risk factor in non COVID-19 mucormycosis, present in 54-76% of such cases in India, Iran, and Mexico^11^. In USA and European countries on the other hand, diabetes mellitus prevalence in patients with mucor mycosis is only 17-23%, whereas haematological malignancies and transplant recipients are the main risk factors reported ^23,24^. In our study, the observed prevalence of diabetes mellitus is 51·9% in CAM. Presence of diabetes mellitus significantly increased the odds of mucormycosis by 4·78 times (95% CI 1·46-15·65). Other studies on CAM from India report a higher prevalence of diabetes (62-80%) presumably related to regional differences in diabetes prevalence ^6,12,18^. New onset diabetes mellitus has been reported more frequently in CAM than in non-CAM cases, this was noted in 21.2% of our patients. New-onset diabetes mellitus in the setting of COVID 19 could be 1) patients with asymptomatic diabetes who had not checked blood sugars recently; 2) patients with pre-diabetes (impaired fasting glucose or impaired glucose tolerance) in whom corticosteroid treatment unmasked diabetes and 3) patients in whom COVID-19 caused pancreatic beta cell dysfunction and reduced insulin production ^15^. Poor glycemic control in known diabetics with CAM (Mean HbA1c 10·08 units, SD 2·7, N=31), could have been due to their inability to consult their regular health-care providers due to lockdown restrictions imposed during the pandemic.

Corticosteroids used in high doses are a known risk factor for mucormycosis ^6,12^, but literature on a specific dose of steroids that confers higher risk is scant. Steroid induced immunosuppression is known to increase the susceptibility to hospital-acquired fungal infections ^25^. Corticosteroids worsen glycemic control and can precipitate ketoacidosis and further increase susceptibility to invasive systemic fungal infections. In our study, 91·6% of subjects were treated with systemic corticosteroids for COVID-19 and a cumulative dexamethasone dose of greater than 120 mg was associated with increased risk of mucormycosis. Even though this dose is higher than the recommended dose of 60 mg dexamethasone over 10 days for severe COVID-19 disease, in many other clinical conditions, higher doses of corticosteroids for longer duration are routinely used. Other Indian studies on CAM report that 76·3-87% of patients were on higher than recommended cumulative dose of corticosteroids^6,18^. Mucormycosis has been reported even after a very short course of corticosteroids for chronic airway disease ^26^. Mucormycosis in our COVID-19 patients occurred after a median time interval of 14 days after diagnosis of COVID-19 and after a median duration of 12 days of corticosteroid therapy, whereas in non-diabetic renal transplant patients, the median time from transplant to mucormycosis was 2·5 months (25). The variability in the steroid dose and duration associated with risk of mucormycosis suggests that corticosteroid use is only one of the factors in the pathogenesis of mucormycosis and that other variables such as the underlying condition may play an important role. COVID-19 is characterized by lymphopenia; other conditions characterized by lymphopenia such as immunosuppressive therapy, cancer chemotherapy, and hematologic malignancies are known risk factors for mucormycosis, therefore it is possible that that COVID-19 per se may predispose to infections such as mucormycosis.

COVID-19 has been well documented to increase risk of thrombosis ^28^. Mucor is a saprophytic mould; it is conceivable that the ischemia and consequent tissue necrosis due to microvascular thrombosis in COVID-19, may be an important cause for invasive mucor infection in COVID 19. Our finding that aspirin and anticoagulation are associated with a reduced risk of mucormycosis supports this contention. We hypothesize that the apparent protective effect of anticoagulation and aspirin against invasive mucor infection may be due to reduction in ischemic tissue necrosis. This hypothesis needs to be tested in additional studies.

## Limitations

There are several limitations to our study. Though case-control study design is best suited to study a rare outcome such as CAM, potential bias in selection of cases and controls may pose threat to validity of our findings. In particular, controls were selected from a single hospital whereas cases were treated in multiple hospitals for COVID-19 before referral to this hospital for treatment of mucormycosis. However, a standardized protocol for treatment of COVID-19 consistent with national and international guidelines is followed in our centre. Our centre is one of the largest private hospitals in the region for treatment of COVID-19 and has patients drawn from all socio-economic strata referred from other hospitals in this region. Therefore, controls drawn from this population may be expected to broadly represent demographic and treatment characteristics of COVID-19 patients from this region. We ensured that the selected controls were hospitalized at approximately the same time as the mucormycosis cases thereby minimizing potential biases arising from changes over time. We were unable to obtain details of pre-admission diabetes control in both groups and have not included details regarding source of oxygen supply or oxygen delivery systems used, factors that can influence infection rates. However, the WHO severity score did not differ in the two groups (Table 2) and the proportion of patients who required oxygen was not different between the two groups suggesting that neither oxygen requirement nor requirement for non-invasive or invasive ventilation were independently associated with mucormycosis. It is possible that the newer variants of concern such as the delta variant may be associated with higher risk of invasive fungal infections but we do not have data regarding genetic sequence of the SARS-CoV-2 virus in our cases and controls.

## Conclusion

Diabetes mellitus and cumulative dexamethasone dose greater than 120 mg (or an equivalent dose of other corticosteroid) are associated with an increased risk of Covid-19 associated mucormycosis. Good glycemic control and limiting cumulative dose of corticosteroids to less than 120 mg of dexamethasone equivalent are likely to reduce risk of CAM. Our observation that use of anticoagulation and aspirin are associated with reduced risk of invasive mucor infection is intriguing and needs further studies. Further research is also needed to understand if COVID-19 itself or a specific genetic variant of SARS-COV2 virus compromises host defence and increases risk of mucormycosis.

## Data Availability

Data Available for review on request

## Bibliography

1. Timeline: WHO’s COVID-19 response [Internet]. [cited 2021 Jul 4]. Available from: https://www.who.int/emergencies/diseases/novel-coronavirus-2019/interactive-timeline

2. Dong E, Du H, Gardner L. An interactive web-based dashboard to track COVID-19 in real time. Lancet Infect Dis. 2020 May;20(5):533–4.

3. Ranjan R, Sharma A, Verma MK. Characterization of the Second Wave of COVID-19 in India. medRxiv. 2021 Apr 21;2021.04.17.21255665.

4. Rawson TM, Moore LSP, Zhu N, Ranganathan N, Skolimowska K, Gilchrist M, et al. Bacterial and fungal co-infection in individuals with coronavirus: A rapid review to support COVID-19 antimicrobial prescribing. Clin Infect Dis. 2020 May 2;ciaa530.

5. Garg D, Muthu V, Sehgal IS, Ramachandran R, Kaur H, Bhalla A, et al. Coronavirus Disease (Covid-19) Associated Mucormycosis (CAM): Case Report and Systematic Review of Literature. Mycopathologia. 2021 May;186(2):289–98.

6. Singh AK, Singh R, Joshi SR, Misra A. Mucormycosis in COVID-19: A systematic review of cases reported worldwide and in India. Diabetes Metab Syndr. 2021;15(4):102146.

7. Sarkar S, Gokhale T, Choudhury SS, Deb AK. COVID-19 and orbital mucormycosis. Indian J Ophthalmol. 2021 Apr;69(4):1002–4.

8. COVID-19 triggering mucormycosis in a susceptible patient: a new phenomenon in the developing world? | BMJ Case Reports [Internet]. [cited 2021 Jul 3]. Available from: https://casereports.bmj.com/content/14/4/e241663

9. Raut A, Huy NT. Rising incidence of mucormycosis in patients with COVID-19: another challenge for India amidst the second wave? The Lancet Respiratory Medicine [Internet]. 2021 Jun 3 [cited 2021 Jul 3];0(0). Available from: https://www.thelancet.com/journals/lanres/article/PIIS2213-2600(21)00265-4/abstract

10. Chakrabarti A. The recent mucormycosis storm over Indian sky. Indian J Med Microbiol. 2021 Jun 23;S0255-0857(21)04134-7.

11. Prakash H, Chakrabarti A. Global Epidemiology of Mucormycosis. J Fungi (Basel). 2019 Mar 21;5(1):26.

12. Patel A, Kaur H, Xess I, Michael JS, Savio J, Rudramurthy S, et al. A multicentre observational study on the epidemiology, risk factors, management and outcomes of mucormycosis in India. Clin Microbiol Infect. 2020 Jul;26(7):944.e9-944.e15.

13. Lin X, Xu Y, Pan X, Xu J, Ding Y, Sun X, et al. Global, regional, and national burden and trend of diabetes in 195 countries and territories: an analysis from 1990 to 2025. Sci Rep. 2020 Sep 8;10(1):14790.

14. RECOVERY Collaborative Group, Horby P, Lim WS, Emberson JR, Mafham M, Bell JL, et al. Dexamethasone in Hospitalized Patients with Covid-19. N Engl J Med. 2021 Feb 25;384(8):693–704.

15. Rubino F, Amiel SA, Zimmet P, Alberti G, Bornstein S, Eckel RH, et al. New-Onset Diabetes in Covid-19. N Engl J Med. 2020 Aug 20;383(8):789–90.

16. John TM, Jacob CN, Kontoyiannis DP. When Uncontrolled Diabetes Mellitus and Severe COVID-19 Converge: The Perfect Storm for Mucormycosis. J Fungi (Basel). 2021 Apr 15;7(4):298.

17. Prakash H, Chakrabarti A. Global Epidemiology of Mucormycosis. J Fungi (Basel). 2019; 5 (1): 26. - Google Search [Internet]. [cited 2021 Jul 3]. Available from: https://www.google.com/search?q=Prakash+H%2C+Chakrabarti+A.+Global+Epidemiology+of+Mucormycosis.+J+Fungi+(Basel).+2019%3B+5+(1)%3A+26.&oq=Prakash+H%2C+Chakrabarti+A.+Global+Epidemiology+of+Mucormycosis.+J+Fungi+(Basel).+2019%3B+5+(1)%3A+26.&aqs=chrome..69i57.2800j0j4&sourceid=chrome&ie=UTF-8

18. Sen M, Honavar SG, Bansal R, Sengupta S, Rao R, Kim U, et al. Epidemiology, clinical profile, management, and outcome of COVID-19-associated rhino-orbital-cerebral mucormycosis in 2826 patients in India - Collaborative OPAI-IJO Study on Mucormycosis in COVID-19 (COSMIC), Report 1. Indian J Ophthalmol. 2021 Jul;69(7):1670–92.

19. Maharashtra records 1,112 mucormycosis deaths, 9,268 cases [Internet]. Free Press Journal. [cited 2021 Jul 21]. Available from: https://www.freepressjournal.in/mumbai/maharashtra-records-1112-mucormycosis-deaths-9268-cases

20. Carfì A, Bernabei R, Landi F, Gemelli Against COVID-19 Post-Acute Care Study Group. Persistent Symptoms in Patients After Acute COVID-19. JAMA. 2020 Aug 11;324(6):603–5.

21. Chopra V, Flanders SA, O’Malley M, Malani AN, Prescott HC. Sixty-Day Outcomes Among Patients Hospitalized With COVID-19. Ann Intern Med. 2021 Apr;174(4):576– 8.

22. Clinical Spectrum [Internet]. COVID-19 Treatment Guidelines. [cited 2021 Jul 4]. Available from: https://www.covid19treatmentguidelines.nih.gov/overview/clinical-spectrum/

23. Skiada A, Pagano L, Groll A, Zimmerli S, Dupont B, Lagrou K, et al. Zygomycosis in Europe: analysis of 230 cases accrued by the registry of the European Confederation of Medical Mycology (ECMM) Working Group on Zygomycosis between 2005 and 2007. Clin Microbiol Infect. 2011 Dec;17(12):1859–67.

24. Kontoyiannis DP, Yang H, Song J, Kelkar SS, Yang X, Azie N, et al. Prevalence, clinical and economic burden of mucormycosis-related hospitalizations in the United States: a retrospective study. BMC Infect Dis. 2016 Dec 1;16(1):730.

25. Lionakis MS, Kontoyiannis DP. Glucocorticoids and invasive fungal infections. Lancet. 2003 Nov 29;362(9398):1828–38.

26. Ferguson AD. Rhinocerebral mucormycosis acquired after a short course of prednisone therapy. J Am Osteopath Assoc. 2007 Nov;107(11):491–3.

27. Song Y, Qiao J, Giovanni G, Liu G, Yang H, Wu J, et al. Mucormycosis in renal transplant recipients: review of 174 reported cases. BMC Infect Dis. 2017 Apr 18;17:283.

28. Levi M, Thachil J, Iba T, Levy JH. Coagulation abnormalities and thrombosis in patients with COVID-19. Lancet Haematol. 2020 Jun;7(6):e438–40.

